# A Cross-Sectional Analysis of Depression and Anxiety Symptoms in the US National Health Interview Survey 2022: A Descriptive Focus on the Black‑Latiné Community

**DOI:** 10.64898/2026.09.23.26361644

**Authors:** Estrella D. Allen, Erendira C. Di Giuseppe, Maria Cruz, Samantha Christie, Ekland Abdiwahab, Kala M. Mehta

**Affiliations:** Department of Chemistry and Biochemistry, San Francisco State University, San Francisco, California, USA; Department of Environmental Health Sciences, Columbia University Mailman School of Public Health, New York, New York, USA; Department of Public Health Sciences, University of California, Davis, Davis, California, USA; Department of Public Health, San Francisco State University, San Francisco, California, USA; Department of Biology, San Francisco State University, San Francisco, California, USA; Department of Epidemiology and Biostatistics, University of California, San Francisco, San Francisco, California, USA

**Keywords:** Black, Latiné, Depression, Anxiety

## Abstract

**Objectives:** To describe the prevalence of anxiety and depression symptoms among Black-Latinés compared to other racial/ethnic groups in the United States.

**Methods:** This cross-sectional analysis leveraged data from the 2022 National Health Interview Survey (NHIS) to estimate the prevalence of anxiety and depression as measured by the GAD-7 and PHQ-8, respectively. Anxiety and depression scores in Black-Latiné were compared to White-Latiné, Non-Latiné White, and Non-Latiné Black respondents.

**Results:** Black-Latinés were more likely to score higher on both the GAD-7 and PHQ-8 indicating higher prevalence of more than minimal anxiety (22.0%) and more than minimal depression (26.0%) than both Non-Latiné Black, anxiety (18.0%) and depression (22.0%), and White-Latiné, anxiety (16.0%) and depression (18.0%), respondents.

**Conclusions:** Black-Latinés are more likely to report more depression and anxiety symptoms than their Non-Latiné Black and White-Latiné counterparts. Studies should try to disaggregate Black subgroups in order to identify the most vulnerable populations.

## Introduction

Health outcomes are often measured in relation to broad racial categories, however, the continued aggregation of health data based on race without consideration to ethnicity, immigration history, and socioeconomic differences not only masks disparities within racial groups but consequently perpetuates existing racial inequalities. Classifying populations based on either race or ethnicity alone may render public health interventions ineffective [1,2]. Despite best attempts, the United States (U.S.) Census Bureau has acknowledged limitations in measuring race and ethnicity [2]. Although race is a social construct and a subjective phenotypic descriptor, research consistently shows that race-a proxy for exposure to racism-is a predictor of health status [3-5]. Unlike race, very little has been done to understand the health effects of the intersection of race and ethnicity [6]. Groups who identify with multiple categories (e.g. Black and Latiné) may possess unique differences compared to those who only identify with one of these categories. This holds for the heterogenous Latiné communities within the U.S. and specifically the Black-Latiné subpopulation [7]. Exploring disaggregated combinations of race and ethnic categories is a pivotal step toward understanding factors that influence adverse health outcomes in previously combined groups [8].

The U.S. and Latin America both codified colorism and racial hierarchy in their national policies [9,10]. In the U.S., state legislation classified individuals based on the “one-drop rule” whereas in Latin America racial classifications were based on how an individual was perceived phenotypically [9,10]. In this study we use the term Latiné instead of Latino(a). The concept of “Latiné” is an inclusive gender-neutral expression more commonly used in the U.S. rather than Latin America to describe individuals with Latin American ancestry or Hispanic origins [11,12]. Although Latin Americans tend to self-identify by referencing their nationality, Latinés demonstrate strong cultural continuity across generations despite their origins [8,13]. Within the broad category of Latiné, we emphasize Black-Latinés which is comprised of individuals of African ancestry from a Latin country and/or mixed individuals with Black and Latiné ancestry. However, both identify as racially Black and ethnically Latiné [8]. Black-Latinés share their racial identity with Non-Latiné Blacks and experience anti-Black racism while also sharing cultural characteristics with White-Latinés within the United States [14,15]. Compared to White-Latinés, Black-Latinés are more likely to be foreign-born and possess unique disadvantages derived from the intersections of race, ethnicity, citizenship, and culture [14].

The global historical impact of racism affects Black-Latinés and is strongly and consistently associated with a range of socioeconomic disadvantages [8]. Compared to White-Latinés, Black-Latinés are more likely to live in poverty (46.2%), to work in service occupations (28.3%), and to receive supplemental nutrition assistance program benefits (27% vs 18%) (SNAP) [14,16,17]. Low socioeconomic status is consistently associated with higher rates of anxiety and depression [18]. Both anxiety and depression have been found to increase chronic disease and consequently higher rates of mortality [19]. Therefore, identifying and addressing poor mental health in this population will have significant effects on chronic conditions and mortality in this population. Being Black and Latiné creates unique health risks; while anti-Black racism negatively impacts health, the Latino paradox which demonstrates that Latiné immigrants have better outcomes than Non-Latiné Blacks and Non-Latiné Whites, confers advantages [9, 20,21]. Due to the complexity of this intersectionality there should be more effort to better understand the prevalence of mental health among Black-Latinés and to tailor mental health interventions to the unique needs of this population [14].

In this study we aim to (1) identify sociodemographic differences between Black-Latinés compared to White-Latinés, Non-Latiné Blacks, and Non-Latiné Whites in the U.S. and (2) to determine the prevalence of depression and anxiety among Black-Latinés compared to White-Latinés, Non-Latiné Blacks, and Non-Latiné Whites in the U.S.

## Methods

### Data Source

For this study, we employed the 2022 National Health Interview Survey (NHIS) [22]. The NHIS, an annual nationally representative survey implemented by the Centers for Disease Control and Prevention (CDC), samples non-institutionalized populations within the U.S. NHIS collects data on health conditions, healthcare access, and mental health outcomes and has been previously used to explore mental health outcomes and disparities within the U.S. [23]. The data that support the findings of this study are available in National Health Interview Survey at https://www.cdc.gov/nchs/nhis/documentation/2022-nhis.html.

### Inclusion/Exclusion Criteria

For this study, analysis was restricted to respondents 18 years of age or older, who completed the Hispanic ethnicity and race questions, and for whom we could calculate anxiety and depression scores using the GAD-7 and PHQ-8.

### Measures

#### i. Disaggregated Race Ethnicity

The NHIS 2022 survey has separate questions for race (single vs. multiple race groups) and ethnicity (Hispanic vs. Non-Hispanic). If Hispanic respondents did not identify with one of the race categories available, the response was coded as “not ascertained,” otherwise this variable was imputed by the NHIS when race was unknown [22]. The available race categories were “White only,” “Black/African American only,” “Asian only,” “American Indian/Alaskan Native only (AIAN),” “AIAN and any other group,” “Other single and multiple races,” refused, not ascertained, and don’t know. Due to possible misclassification, our analysis dropped individuals who selected “Other single and multiple races,” refused, not ascertained, and don’t know. For our analysis, we restricted to individuals who identified racially as either Black/African American only, White only, those who selected Black/African American and Hispanic identity, and those who selected White race and Hispanic identity. We generated a new race/ethnicity variable with the following categories: Black-Latiné (Black and Hispanic), White-Latiné (White and Hispanic), Non-Latiné Black (Black and Non-Hispanic), and White Non-Latiné (White and Non-Hispanic).

#### ii. Mental Health Symptoms

Anxiety was measured using the validated and widely used General Anxiety Disorder-7 (GAD-7) [22]. The GAD-7 gathers anxiety symptoms over the last two weeks using 7 questions: the total sum of all questions per person ranges from 0 to 21. The resulting total was sorted into the following categories: minimal (0-4), mild (5-9), moderate (10-14), and severe anxiety (15-21). Similarly, depression was measured using the validated and widely used Patient Health Questionnaire-8 (PHQ-8) that also captures depression symptoms over the past two weeks using 8 questions [22]. The sum of all questions per person ranges from 0 to 24 and respondents were categorized as having none/minimal (0-4), mild (5-9), moderate (10-14), and severe (15-24) depressed. In addition, to determine the prevalence of clinically diagnosed anxiety and depression, respondents were asked if a physician ever diagnosed them with any anxiety disorder or depression.

#### iii. Demographics

The following demographic variables were summarized: age (years); sex (male/female), and highest education (less than high school, high school graduate, secondary college or associate’s degree, college degree or higher); marital status (married/living with a partner or not married/living with a partner); sexual orientation (straight, bisexual, gay/lesbian, something else); and has health insurance (yes/no). Respondents were also asked if they were born in the United States and/or had U.S. citizenship (yes/no). Respondents who reported they were not born in the U.S. were asked to provide the number of years they lived in the U.S.: “Less than 1 year,” “1 to 5 years,” “5 to less than 10 years,” “10 to less than 15 years,” and “15 years or more.” We collapsed this response into three categories, “less than 5 years,” “5 to 10 years,” and “10 years or more” since the immigrant effect has been associated with those time intervals [24].

Respondents’ socioeconomic status was determined by how many hours they work per week (≥35 hours per week vs. <35 hours per week) and by calculating their ratio of income-to-poverty level (continuous) [25,26]. Values <1 indicate total pre-tax income below the federal poverty level, a value of 2 means the total pre-tax income was double the federal poverty threshold for that year in question.

### Analysis

Univariate and bivariate analysis including frequency tables, chi-squared test with Rao & Scott’s second order correction, and Wilcoxon rank-sum test for complex survey samples were employed to describe sample characteristics and to estimate the prevalence of anxiety and depression within our racial/ethnic subgroups.

Statistical significance was set at p < 0.05 and all analysis was performed using Rstudio [27].

### Ethical Approval

This research is exempt certification based on UCSF IRB under category 4: secondary research for which consent is not required. IRB Number: 25-44668

## Results

Table 1 presents both the weighted and unweighted demographic characteristics of the analytic sample. In general, the sample population was Non-Latiné White (74%), married/living with a partner (60.8%), heterosexual (94%), had at least a secondary/associate’s degree (63%), worked ≥35 hours per week (81%), had medical insurance (93%), was born in the U.S. (89%), and had U.S. citizenship (96%). Table 2 presents the demographic differences between the racial/ethnic subgroups. Black-Latiné respondents were mainly female (53%) and tended to be younger than the comparison groups; median age was 36 years. Black-Latinés were less likely than both White-Latinés (61%) and Non-Latiné Whites (64%) to be married or Living with a partner (43%) and were less educated than comparison groups with only 16% of respondents attaining a college degree or higher. Black-Latinés were more likely to have a statistically significant lower income-to-poverty ratio compared to other groups (p<.001). Compared to other groups, Black-Latinés were more likely to be recent immigrants with 20% of non-US born respondents reporting being in the U.S. less than 5 years. Figures 1 and 2 show the survey-weighted distribution of anxiety symptoms (GAD-7) and depression symptoms (PHQ-8), respectively. We also plotted the cutoffs used when scoring these measures, and found that ~80% of those surveyed fell under the “none/minimal” category.

**Table 1.** Sample characteristics (weighted and unweighted)

| **Characteristic<sup>a</sup>** | **Weighted**<br>**N = 214,530,725** | **Unweighted**<br>**N = 23,705** |
| --- | --- | --- |
| **Age** | 49 (33, 64) | 56 (38, 69) |
| **Sex** |  |  |
| Female | 110,165,685 (51%) | 12,927 (55%) |
| Male | 104,337,781 (49%) | 10,775 (45%) |
| **Race/Ethnicity** |  |  |
| Black-Latiné | 1,243,567 (0.6%) | 119 (0.5%) |
| Non-Latiné Black | 30,339,270 (14%) | 3,112 (13%) |
| White-Latiné | 24,452,592 (11%) | 2,232 (9.4%) |
| Non-Latiné White | 158,495,296 (74%) | 18,242 (77%) |
| **Marital status** |  |  |
| Married/living with a partner | 124,139,699 (60.8%) | 11,932 (52.4%) |
| Neither | 80,788,400 (39.2%) | 10,781 (47.6%) |
| **Sexual orientation** |  |  |
| Straight | 190,322,214 (94%) | 21,248 (95%) |
| Gay/Lesbian | 4,242,166 (2.1%) | 500 (2.2%) |
| Bisexual | 5,486,236 (2.7%) | 484 (2.2%) |
| Something else | 1,698,179 (0.8%) | 156 (0.7%) |
| **Highest level of education attained** |  |  |
| Less than high school | 19,707,777 (9.2%) | 1,784 (7.6%) |
| High school graduate | 58,346,811 (27%) | 6,057 (26%) |
| Secondary/Associate's degree | 64,751,049 (30%) | 6,829 (29%) |
| College degree or higher | 70,617,245 (33%) | 8,938 (38%) |
| **Income-to-poverty ratio** | 11 (7, 14) | 11 (7, 14) |
| **Employed 35+ hours/week** | 103,792,023 (81%) | 10,345 (81%) |
| **Has any medical insurance** | 198,845,440 (93%) | 22,342 (94%) |
| **Born in the United States** | 181,592,609 (89%) | 20,438 (90%) |
| **Has United States citizenship** | 195,151,033 (96%) | 21,879 (97%) |
| **If not born in the US, years in the United States** |  |  |
| Less than 5 years | 1,758,368 (7.9%) | 143 (6.5%) |
| 5 to 15 years | 4,657,841 (21%) | 404 (18%) |
| 15 years or more | 15,919,944 (71%) | 1,637 (75%) |

<sup>a</sup>Median (IQR); n (%).

**Table 2.** Weighted sample characteristics by race and ethnicity.

| Characteristic<sup>a</sup> | Race/Ethnicity |  |  |  | p-value<sup>b</sup> |
| --- | --- | --- | --- | --- | --- |
|  | Black-Latiné<br>N = 1,243,567 | Non-Latiné Black<br>N = 30,339,270 | White-Latiné<br>N = 24,452,592 | Non-Latiné White<br>N = 158,495,296 |  |
| **Age** | 36.0 (25.0, 51.0) | 45.0 (31.0, 60.0) | 41.0 (29.0, 55.0) | 51.0 (34.0, 66.0) | <0.001 |
| **Sex** |  |  |  |  | <0.001 |
| Female | 53.0% | 56.0% | 52.0% | 50.0% |  |
| Male | 47.0% | 44.0% | 48.0% | 50.0% |  |
| **Marital status** |  |  |  |  | <0.001 |
| Married/living with a partner | 43.0% | 40.7% | 61.0% | 64.0% |  |
| Neither | 57.0% | 59.3% | 39.0% | 36.0% |  |
| **Sexual orientation** |  |  |  |  | 0.6 |
| Straight | 97.0% | 94.8% | 95.0% | 94.1% |  |
| Gay/Lesbian | 0.0% | 1.8% | 1.9% | 2.2% |  |
| Bisexual | 2.1% | 2.6% | 2.4% | 2.8% |  |
| Something else | 0.9% | 0.8% | 0.7% | 0.9% |  |
| **Highest level of education attained** |  |  |  |  | <0.001 |
| Less than high school | 25.0% | 13.0% | 23.0% | 7.0% |  |
| High school graduate | 32.0% | 33.0% | 29.0% | 26.0% |  |
| Secondary/Associate's degree | 27.0% | 31.0% | 29.0% | 30.0% |  |
| College degree or higher | 16.0% | 23.0% | 19.0% | 37.0% |  |
| **Income-to-poverty ratio** | 8.0 (4.0, 10.0) | 8.0 (5.0, 12.0) | 8.0 (5.0, 12.0) | 12.0 (8.0, 14.0) | <0.001 |
| **Employed 35+ hours/week** | 79.0% | 80.0% | 79.0% | 81.0% | 0.3 |
| **Has any medical insurance** | 91.0% | 90.0% | 80.0% | 95.0% | <0.001 |
| **Born in the United States** | 66.0% | 85.0% | 54.0% | 95.0% | <0.001 |
| **Has United States citizenship** | 84.0% | 95.0% | 75.0% | 99.0% | <0.001 |
| **If not born in the US, years in the United States** |  |  |  |  | <0.001 |
| Less than 5 years | 20.0% | 8.5% | 8.5% | 6.0% |  |
| 5 to 15 years | 23.0% | 31.0% | 20.0% | 17.0% |  |
| 15 years or more | 57.0% | 61.0% | 72.0% | 77.0% |  |

<sup>a</sup>Median (IQR); %.<sup>b</sup>Chi-squared test with Rao & Scott's second-order correction; Wilcoxon rank-sum test for complex survey samples.

**Figure 1.**
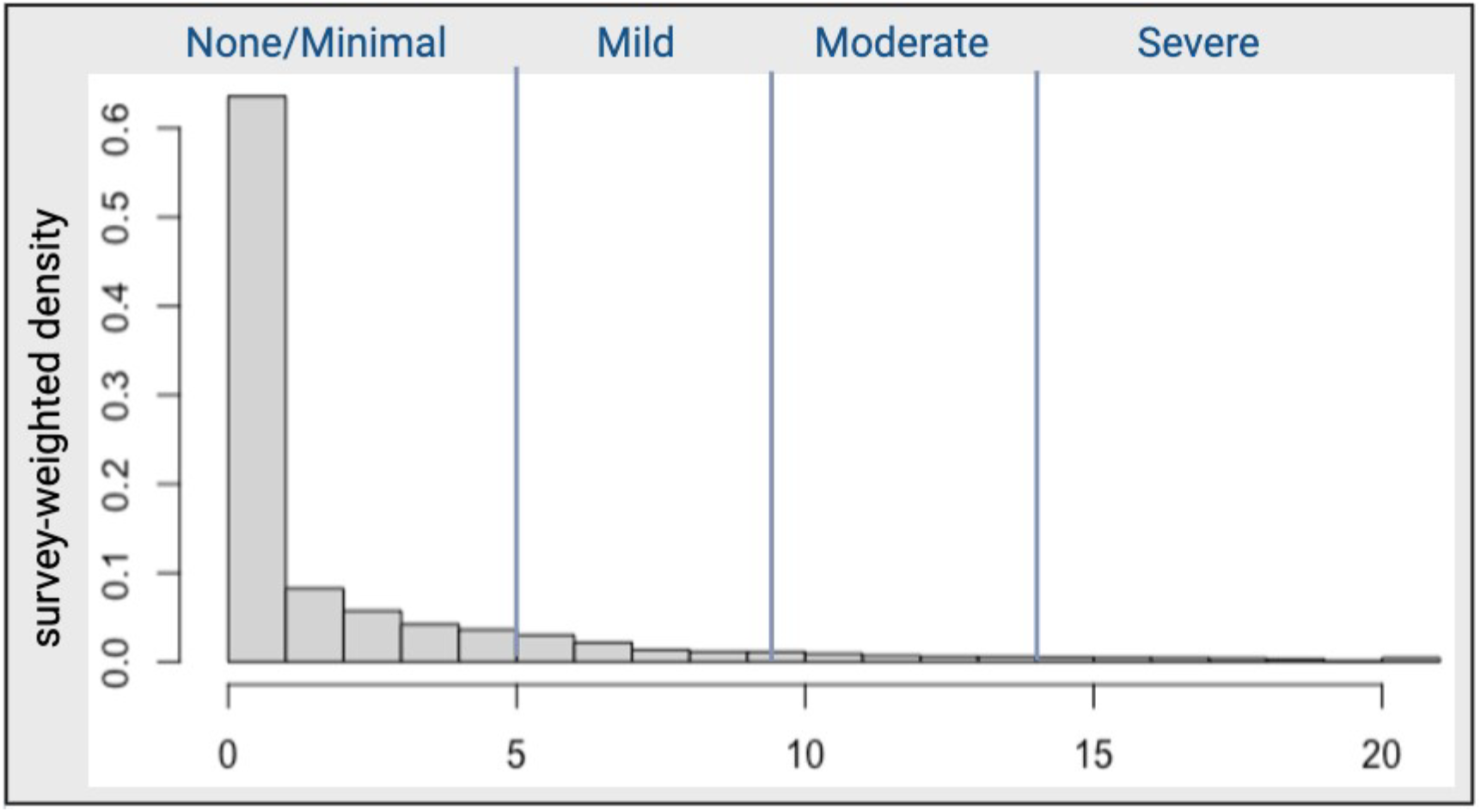
Distribution of total GAD-7 scores and scale categorization for entire study sample.

**Figure 2.**
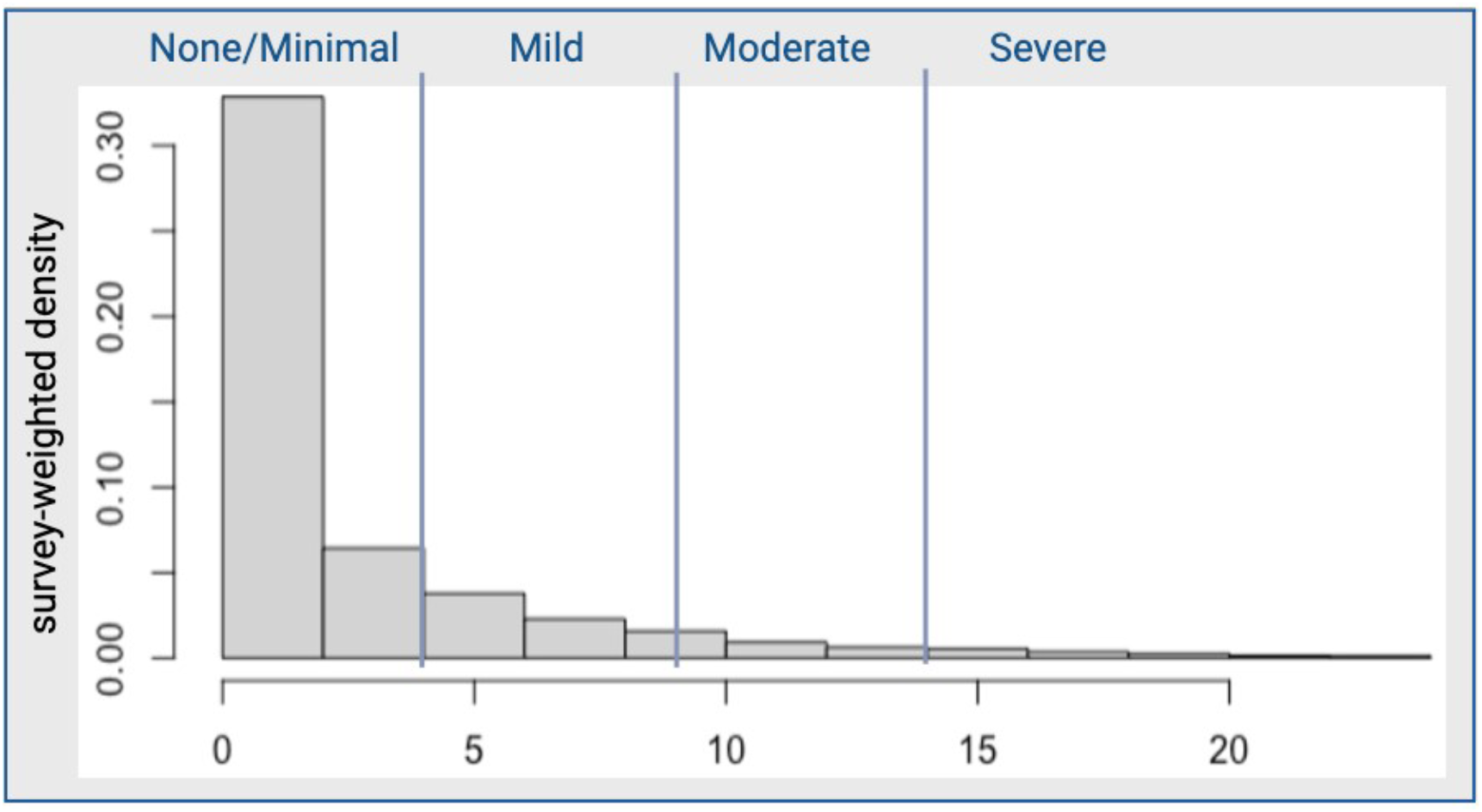
Distribution of total PHQ-8 scores and scale categorization for entire study sample.

The prevalence of anxiety symptoms differed by population subgroups (Table 3, Figure 3). Black-Latinés were more likely to report a medical history of anxiety than both Non-Latiné Blacks and White-Latinés, with 17% reporting they were told by a medical physician or other medical professional that they had an anxiety disorder. Black-Latinés were more likely to report “feeling nervous, anxiety, or on edge,” and “becoming easily annoyed or irritable” more than half of the days. Black-Latinés were also more likely to report experiencing “worrying too much about different things” (6.8%) and “trouble relaxing” (6.9%) nearly every day. Based on the GAD-7, Black-Latinés were more likely than other groups to be categorized with more than minimal anxiety (22.0%). That said, there was no statistically significant difference in GAD-7 anxiety categories between race/ethnicity groups (p=0.30).

**Table 3.**
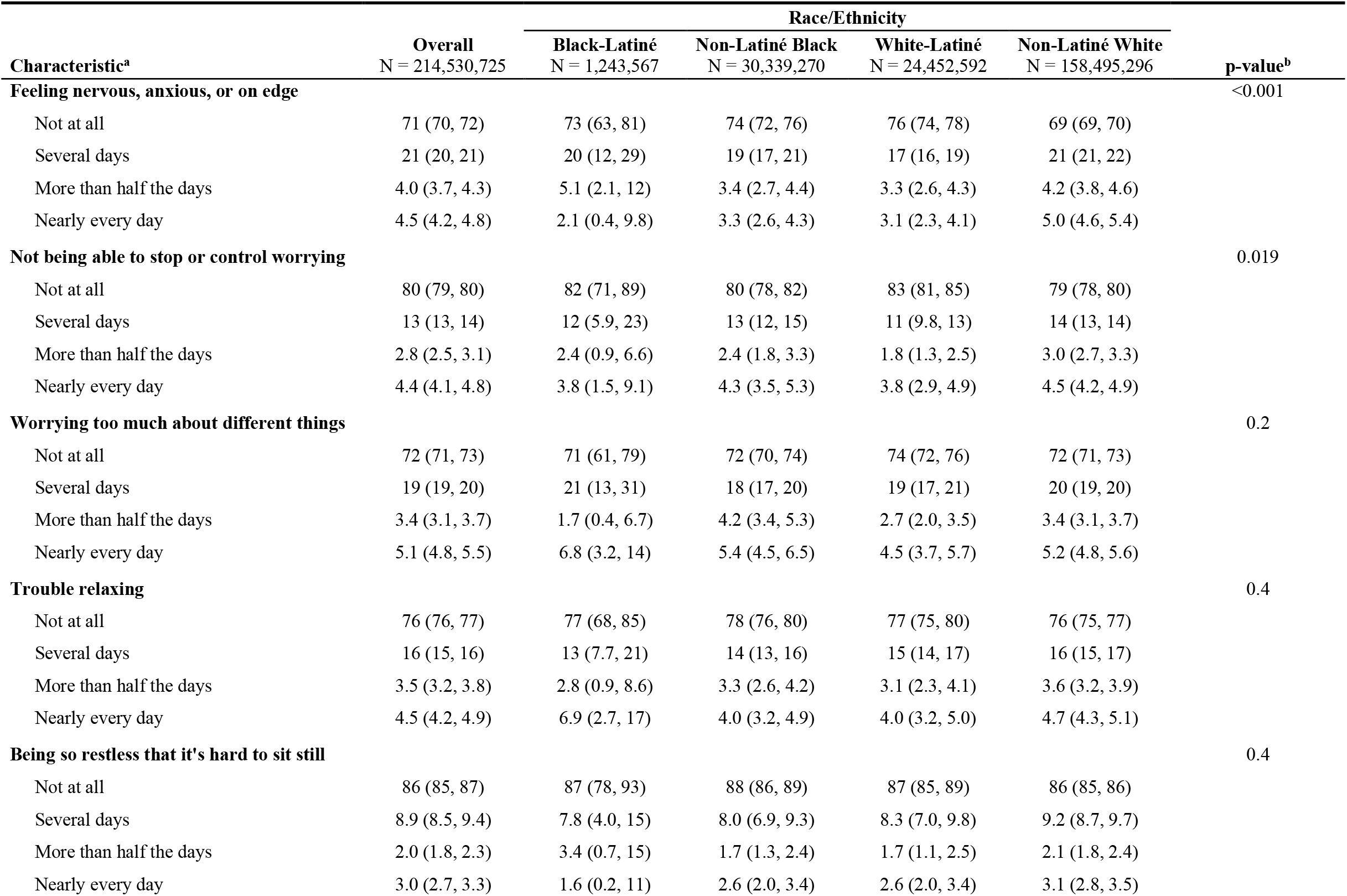
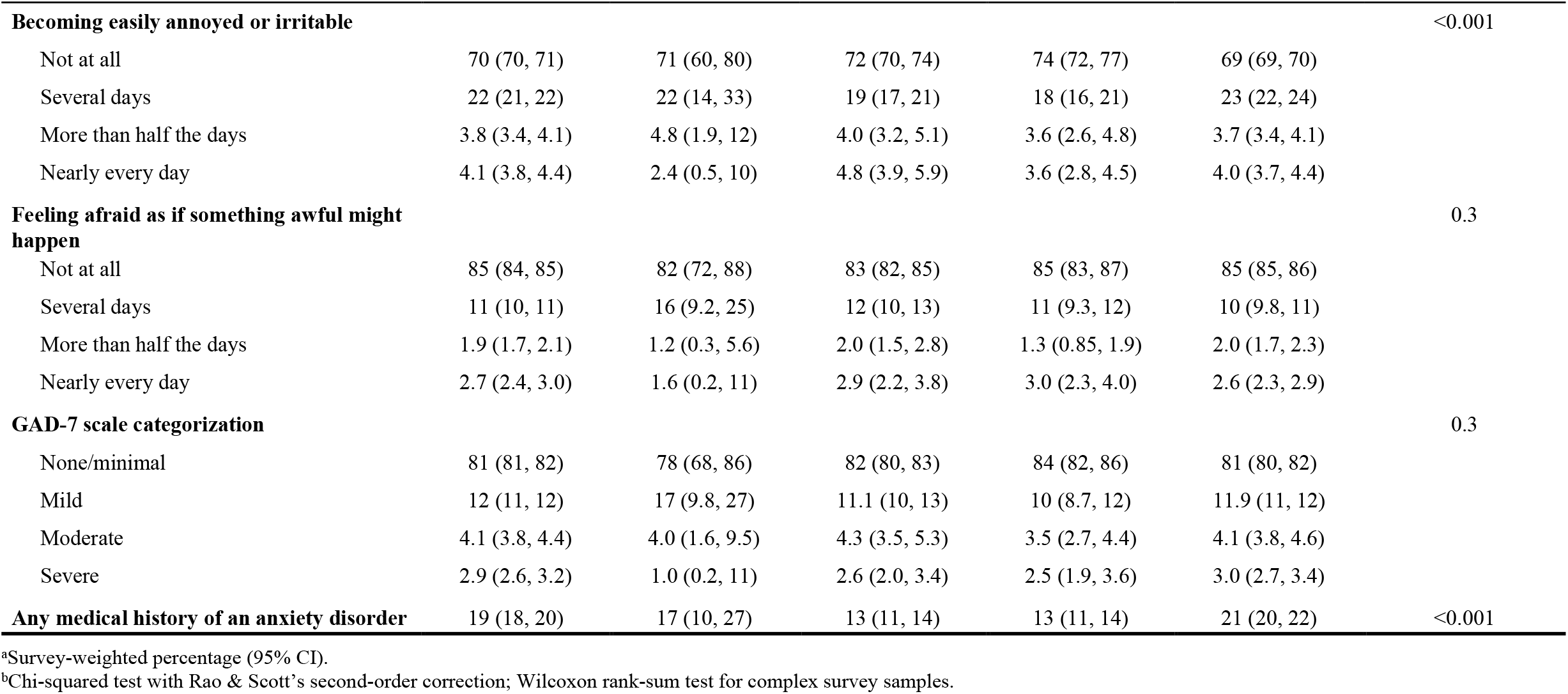
GAD-7 anxiety symptoms by race and ethnicity.

| Characteristic<sup>a</sup> | Overall<br>N = 214,530,725 | Race/Ethnicity |  |  |  | p-value<sup>b</sup> |
| --- | --- | --- | --- | --- | --- | --- |
|  |  | Black-Latiné<br>N = 1,243,567 | Non-Latiné Black<br>N = 30,339,270 | White-Latiné<br>N = 24,452,592 | Non-Latiné White<br>N = 158,495,296 |  |
| **Feeling nervous, anxious, or on edge** |  |  |  |  |  | <0.001 |
| Not at all | 71 (70, 72) | 73 (63, 81) | 74 (72, 76) | 76 (74, 78) | 69 (69, 70) |  |
| Several days | 21 (20, 21) | 20 (12, 29) | 19 (17, 21) | 17 (16, 19) | 21 (21, 22) |  |
| More than half the days | 4.0 (3.7, 4.3) | 5.1 (2.1, 12) | 3.4 (2.7, 4.4) | 3.3 (2.6, 4.3) | 4.2 (3.8, 4.6) |  |
| Nearly every day | 4.5 (4.2, 4.8) | 2.1 (0.4, 9.8) | 3.3 (2.6, 4.3) | 3.1 (2.3, 4.1) | 5.0 (4.6, 5.4) |  |
| **Not being able to stop or control worrying** |  |  |  |  |  | 0.019 |
| Not at all | 80 (79, 80) | 82 (71, 89) | 80 (78, 82) | 83 (81, 85) | 79 (78, 80) |  |
| Several days | 13 (13, 14) | 12 (5.9, 23) | 13 (12, 15) | 11 (9.8, 13) | 14 (13, 14) |  |
| More than half the days | 2.8 (2.5, 3.1) | 2.4 (0.9, 6.6) | 2.4 (1.8, 3.3) | 1.8 (1.3, 2.5) | 3.0 (2.7, 3.3) |  |
| Nearly every day | 4.4 (4.1, 4.8) | 3.8 (1.5, 9.1) | 4.3 (3.5, 5.3) | 3.8 (2.9, 4.9) | 4.5 (4.2, 4.9) |  |
| **Worrying too much about different things** |  |  |  |  |  | 0.2 |
| Not at all | 72 (71, 73) | 71 (61, 79) | 72 (70, 74) | 74 (72, 76) | 72 (71, 73) |  |
| Several days | 19 (19, 20) | 21 (13, 31) | 18 (17, 20) | 19 (17, 21) | 20 (19, 20) |  |
| More than half the days | 3.4 (3.1, 3.7) | 1.7 (0.4, 6.7) | 4.2 (3.4, 5.3) | 2.7 (2.0, 3.5) | 3.4 (3.1, 3.7) |  |
| Nearly every day | 5.1 (4.8, 5.5) | 6.8 (3.2, 14) | 5.4 (4.5, 6.5) | 4.5 (3.7, 5.7) | 5.2 (4.8, 5.6) |  |
| **Trouble relaxing** |  |  |  |  |  | 0.4 |
| Not at all | 76 (76, 77) | 77 (68, 85) | 78 (76, 80) | 77 (75, 80) | 76 (75, 77) |  |
| Several days | 16 (15, 16) | 13 (7.7, 21) | 14 (13, 16) | 15 (14, 17) | 16 (15, 17) |  |
| More than half the days | 3.5 (3.2, 3.8) | 2.8 (0.9, 8.6) | 3.3 (2.6, 4.2) | 3.1 (2.3, 4.1) | 3.6 (3.2, 3.9) |  |
| Nearly every day | 4.5 (4.2, 4.9) | 6.9 (2.7, 17) | 4.0 (3.2, 4.9) | 4.0 (3.2, 5.0) | 4.7 (4.3, 5.1) |  |
| **Being so restless that it's hard to sit still** |  |  |  |  |  | 0.4 |
| Not at all | 86 (85, 87) | 87 (78, 93) | 88 (86, 89) | 87 (85, 89) | 86 (85, 86) |  |
| Several days | 8.9 (8.5, 9.4) | 7.8 (4.0, 15) | 8.0 (6.9, 9.3) | 8.3 (7.0, 9.8) | 9.2 (8.7, 9.7) |  |
| More than half the days | 2.0 (1.8, 2.3) | 3.4 (0.7, 15) | 1.7 (1.3, 2.4) | 1.7 (1.1, 2.5) | 2.1 (1.8, 2.4) |  |
| Nearly every day | 3.0 (2.7, 3.3) | 1.6 (0.2, 11) | 2.6 (2.0, 3.4) | 2.6 (2.0, 3.4) | 3.1 (2.8, 3.5) |  |

| Characteristic<sup>a</sup> | Race/Ethnicity |  |  |  |  | p-value<sup>b</sup> |
| --- | --- | --- | --- | --- | --- | --- |
|  | Overall<br>N = 214,530,725 | Black-Latiné<br>N = 1,243,567 | Non-Latiné Black<br>N = 30,339,270 | White-Latiné<br>N = 24,452,592 | Non-Latiné White<br>N = 158,495,296 |  |
| **Becoming easily annoyed or irritable** |  |  |  |  |  | <0.001 |
| Not at all | 70 (70, 71) | 71 (60, 80) | 72 (70, 74) | 74 (72, 77) | 69 (69, 70) |  |
| Several days | 22 (21, 22) | 22 (14, 33) | 19 (17, 21) | 18 (16, 21) | 23 (22, 24) |  |
| More than half the days | 3.8 (3.4, 4.1) | 4.8 (1.9, 12) | 4.0 (3.2, 5.1) | 3.6 (2.6, 4.8) | 3.7 (3.4, 4.1) |  |
| Nearly every day | 4.1 (3.8, 4.4) | 2.4 (0.5, 10) | 4.8 (3.9, 5.9) | 3.6 (2.8, 4.5) | 4.0 (3.7, 4.4) |  |
| **Feeling afraid as if something awful might happen** |  |  |  |  |  | 0.3 |
| Not at all | 85 (84, 85) | 82 (72, 88) | 83 (82, 85) | 85 (83, 87) | 85 (85, 86) |  |
| Several days | 11 (10, 11) | 16 (9.2, 25) | 12 (10, 13) | 11 (9.3, 12) | 10 (9.8, 11) |  |
| More than half the days | 1.9 (1.7, 2.1) | 1.2 (0.3, 5.6) | 2.0 (1.5, 2.8) | 1.3 (0.85, 1.9) | 2.0 (1.7, 2.3) |  |
| Nearly every day | 2.7 (2.4, 3.0) | 1.6 (0.2, 11) | 2.9 (2.2, 3.8) | 3.0 (2.3, 4.0) | 2.6 (2.3, 2.9) |  |
| **GAD-7 scale categorization** |  |  |  |  |  | 0.3 |
| None/minimal | 81 (81, 82) | 78 (68, 86) | 82 (80, 83) | 84 (82, 86) | 81 (80, 82) |  |
| Mild | 12 (11, 12) | 17 (9.8, 27) | 11.1 (10, 13) | 10 (8.7, 12) | 11.9 (11, 12) |  |
| Moderate | 4.1 (3.8, 4.4) | 4.0 (1.6, 9.5) | 4.3 (3.5, 5.3) | 3.5 (2.7, 4.4) | 4.1 (3.8, 4.6) |  |
| Severe | 2.9 (2.6, 3.2) | 1.0 (0.2, 11) | 2.6 (2.0, 3.4) | 2.5 (1.9, 3.6) | 3.0 (2.7, 3.4) |  |
| **Any medical history of an anxiety disorder** | 19 (18, 20) | 17 (10, 27) | 13 (11, 14) | 13 (11, 14) | 21 (20, 22) | <0.001 |

<sup>a</sup>Survey-weighted percentage (95% CI).
<sup>b</sup>Chi-squared test with Rao & Scott's second-order correction; Wilcoxon rank-sum test for complex survey samples.

**Figure 3.**
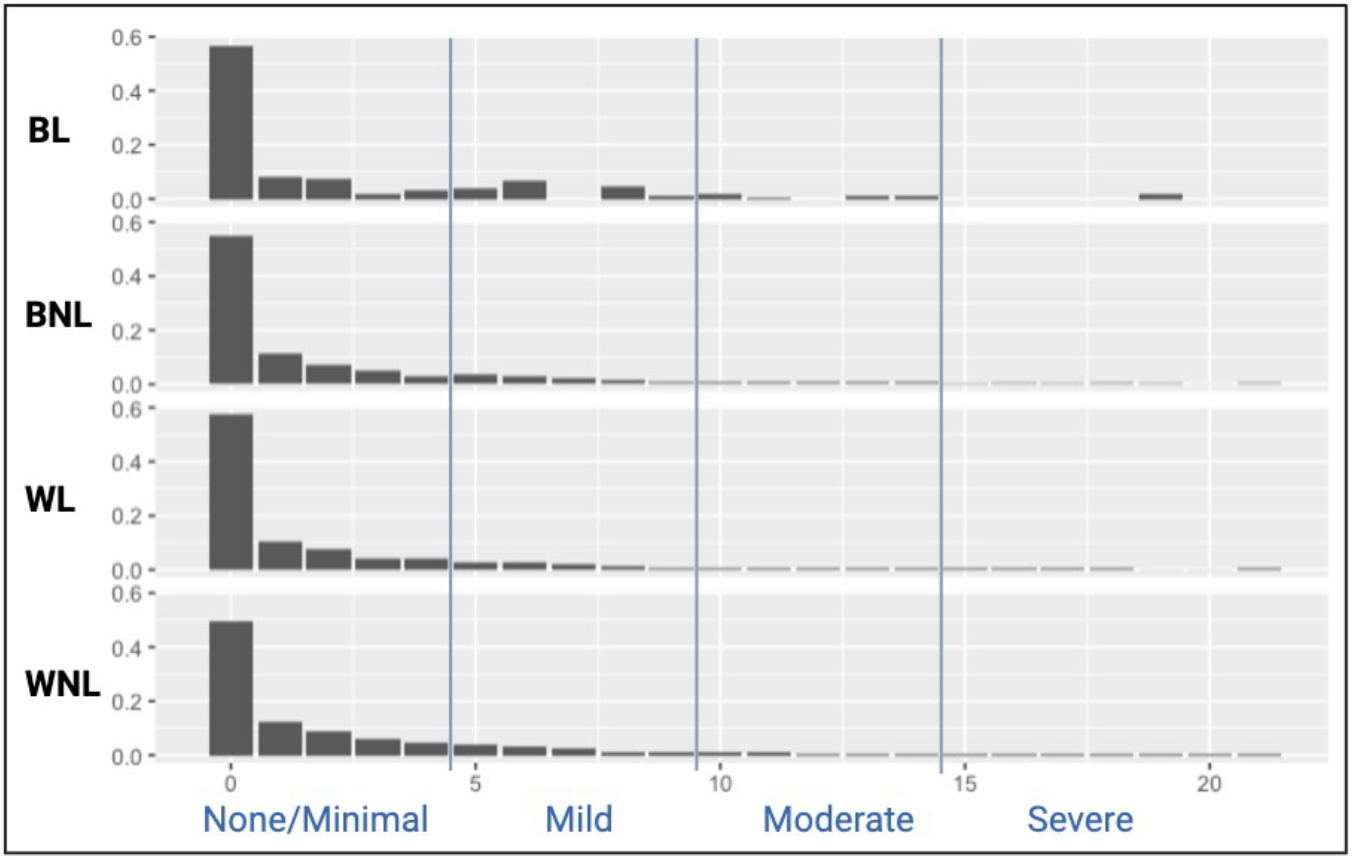
Survey-weighted density distributions of GAD-7 scores and scale categorization by race/ethnicity. BL, Black-Latiné; BNL, Black Non-Latiné; WL, White-Latiné; WNL, White Non-Latiné. Vertical lines mark scale category cutoffs.

The prevalence of depression symptoms also differed by population subgroups (Table 4, Figure 4). Black-Latinés were more likely to report a medical history of depression than both Non-Latiné Blacks and White-Latinés, with 16% reporting they were told by a medical physician or other medical professional that they had depression. Black-Latinés were more likely to report they experienced most of the depression symptoms including “feeling down, depressed, or hopeless”, having “trouble falling asleep or staying asleep, or sleeping too much”, and “feeling tired or having little energy” more than half of the days over the past two weeks. There was a statistically significant difference in the PHQ-8 depression scale between race/ethnicity groups (p<0.001) with Black-Latinés more likely than other groups to be categorized with more than minimal depression (26.2%).

**Table 4.** PHQ-8 depression symptoms by race and ethnicity.

| Characteristic<sup>a</sup> | Overall<br>N = 214,530,725 | Race/Ethnicity |  |  |  | p-value<sup>b</sup> |
| --- | --- | --- | --- | --- | --- | --- |
|  |  | Black-Latiné<br>N = 1,243,567 | Non-Latiné Black<br>N = 30,339,270 | White-Latiné<br>N = 24,452,592 | Non-Latiné White<br>N = 158,495,296 |  |
| **Little interest or pleasure in doing things** |  |  |  |  |  | <0.001 |
| Not at all | 80 (79, 81) | 75 (66, 83) | 77 (75, 79) | 83 (82, 85) | 80 (79, 81) |  |
| Several days | 14 (13, 14) | 19 (13, 29) | 15 (13, 16) | 12 (10, 13) | 14 (13, 14) |  |
| More than half the days | 3.3 (3.0, 3.6) | 2.4 (0.87, 6.6) | 4.2 (3.4, 5.3) | 2.5 (1.8, 3.4) | 3.2 (2.9, 3.5) |  |
| Nearly every day | 3.1 (2.9, 3.4) | 2.7 (0.87, 8.1) | 3.8 (3.0, 4.7) | 2.3 (1.7, 3.1) | 3.1 (2.9, 3.5) |  |
| **Feeling down, depressed, or hopeless** |  |  |  |  |  | 0.2 |
| Not at all | 80 (79, 81) | 76 (66, 84) | 80 (78, 82) | 83 (81, 85) | 80 (79, 81) |  |
| Several days | 15 (14, 15) | 17 (11, 26) | 15 (13, 16) | 12 (11, 14) | 15 (14, 16) |  |
| More than half the days | 2.6 (2.3, 2.8) | 4.2 (1.7, 9.6) | 2.8 (2.2, 3.6) | 2.2 (1.6, 3.1) | 2.5 (2.3, 2.9) |  |
| Nearly every day | 2.7 (2.5, 3.0) | 2.7 (0.7, 9.6) | 2.2 (3.6) | 2.4 (1.7, 3.3) | 2.7 (2.5, 3.0) |  |
| **Trouble falling asleep or staying asleep, or sleeping too much** |  |  |  |  |  | <0.001 |
| Not at all | 64 (63, 65) | 68 (56, 78) | 68 (66, 70) | 69 (67, 72) | 62 (61, 63) |  |
| Several days | 21 (20, 22) | 21 (13, 31) | 19 (17, 21) | 19 (17, 21) | 22 (21, 23) |  |
| More than half the days | 5.4 (5.1, 5.8) | 7.3 (3.1, 16) | 4.4 (3.6, 5.4) | 4.1 (3.1, 5.5) | 5.8 (5.4, 6.2) |  |
| Nearly every day | 9.8 (9.3, 10) | 4.2 (1.4, 12) | 8.8 (7.6, 10) | 8.0 (6.6, 9.8) | 10 (9.7, 11) |  |
| **Feeling tired or having little energy** |  |  |  |  |  | <0.001 |
| Not at all | 56 (55, 57) | 57 (47, 67) | 58 (56, 60) | 62 (59, 64) | 54 (53, 55) |  |
| Several days | 29 (28, 30) | 29 (21, 40) | 28 (26, 30) | 27 (25, 29) | 29 (28, 30) |  |
| More than half the days | 6.1 (5.7, 6.4) | 6.1 (2.9, 12) | 5.7 (4.8, 6.9) | 5.0 (4.1, 6.2) | 6.3 (5.9, 6.7) |  |
| Nearly every day | 9.5 (9.0, 10) | 7.0 (3.4, 14) | 8.4 (7.2, 9.7) | 6.7 (5.6, 8.0) | 10 (9.5, 11) |  |
| **Poor appetite or overeating** |  |  |  |  |  | 0.13 |
| Not at all | 80 (79, 80) | 78 (69, 86) | 77 (76, 79) | 79 (77, 81) | 80 (79, 81) |  |
| Several days | 12 (12, 13) | 12 (7.3, 20) | 14 (13, 16) | 13 (12, 15) | 12 (11, 13) |  |
| More than half the days | 3.2 (3.0, 3.5) | 3.4 (1.5, 7.8) | 3.6 (2.8, 4.5) | 3.5 (2.7, 4.5) | 3.1 (2.9, 3.5) |  |
| Nearly every day | 4.7 (4.3, 5.0) | 5.8 (2.5, 13) | 5.1 (4.3, 6.1) | 3.8 (3.0, 4.8) | 4.7 (4.3, 5.1) |  |

| Characteristic<sup>a</sup> | Race/Ethnicity |  |  |  |  | p-value<sup>b</sup> |
| --- | --- | --- | --- | --- | --- | --- |
|  | Overall<br>N = 214,530,725 | Black-Latiné<br>N = 1,243,567 | Non-Latiné Black<br>N = 30,339,270 | White-Latiné<br>N = 24,452,592 | Non-Latiné White<br>N = 158,495,296 |  |
| **Feeling bad about yourself, that you are a failure, or have let yourself/family down** |  |  |  |  |  | 0.018 |
| Not at all | 85 (84, 85) | 81 (71, 88) | 86 (85, 88) | 86 (84, 88) | 84 (83, 85) |  |
| Several days | 10 (9.8, 11) | 16 (9.0, 26) | 8.8 (7.5, 10) | 8.8 (7.7, 10) | 11 (10, 11) |  |
| More than half the days | 2.2 (2.0, 2.4) | 2.5 (0.9, 6.8) | 1.8 (1.3, 2.4) | 2.4 (1.7, 3.3) | 2.2 (2.0, 2.5) |  |
| Nearly every day | 3.0 (2.7, 3.3) | 0.5 (0.1, 3.8) | 3.0 (2.2, 4.0) | 2.5 (1.8, 3.5) | 3.1 (2.7, 3.4) |  |
| **Trouble concentrating on things, such as reading or watching TV** |  |  |  |  |  | 0.021 |
| Not at all | 84 (83, 85) | 87 (79, 93) | 85 (83, 87) | 86 (85, 88) | 83 (83, 84) |  |
| Several days | 10 (9.8, 11) | 8.5 (4.3, 16) | 9.1 (7.9, 10) | 9.3 (8.1, 11) | 11 (10, 11) |  |
| More than half the days | 2.4 (2.1, 2.6) | 2.7 (0.7, 9.4) | 2.2 (1.6, 3.0) | 2.1 (1.6, 2.9) | 2.4 (2.1, 2.7) |  |
| Nearly every day | 3.4 (3.2, 3.7) | 1.5 (0.4, 6.0) | 3.6 (2.8, 4.6) | 2.2 (1.6, 3.0) | 3.6 (3.3, 4.0) |  |
| **Moving or speaking so slowly, or the opposite, being so fidgety or restless that others have noticed** |  |  |  |  |  | 0.046 |
| Not at all | 93 (92, 93) | 90 (79, 95) | 92 (91, 93) | 92 (90, 93) | 93 (93, 94) |  |
| Several days | 4.5 (4.1, 4.8) | 7.3 (2.7, 18) | 4.8 (4.0, 5.9) | 5.8 (4.8, 7.0) | 4.2 (3.8, 4.6) |  |
| More than half the days | 1.1 (0.9, 1.2) | 3.1 (0.9, 10) | 1.2 (0.8, 1.8) | 0.7 (0.4, 1.1) | 1.1 (0.9, 1.3) |  |
| Nearly every day | 1.7 (1.5, 1.9) | 0.0 (0.0, 0.0) | 1.8 (1.3, 2.6) | 1.7 (1.2, 2.4) | 1.7 (1.5, 1.9) |  |
| **PHQ-8 scale categorization** |  |  |  |  |  | 0.004 |
| None/minimal | 78 (77, 79) | 73.8 (64, 82) | 77.9 (76, 80) | 82 (80, 84) | 78 (77, 78) |  |
| Mild | 14.3 (14, 15) | 20 (13, 29) | 14 (13, 16) | 11 (9.6, 12) | 15 (14, 15) |  |
| Moderate | 4.7 (4.4, 5.1) | 3.1 (1.1, 8.4) | 5.4 (4.4, 6.5) | 4.3 (3.3, 5.2) | 4.0 (4.3, 5.1) |  |
| Severe | 3.0 (2.7, 3.3) | 3.1 (0.9, 10) | 2.7 (2.1, 3.5) | 2.7 (2.0, 3.8) | 3.0 (2.8, 3.4) |  |
| **Any medical history of depression** | 19 (19, 20) | 16 (9.8, 25) | 14 (12, 15) | 14 (12, 15) | 21 (21, 22) | <0.001 |

<sup>a</sup>Survey-weighted percentage (95% CI).
<sup>b</sup>Chi-squared test with Rao & Scott's second-order correction; Wilcoxon rank-sum test for complex survey samples.

**Figure 4.**
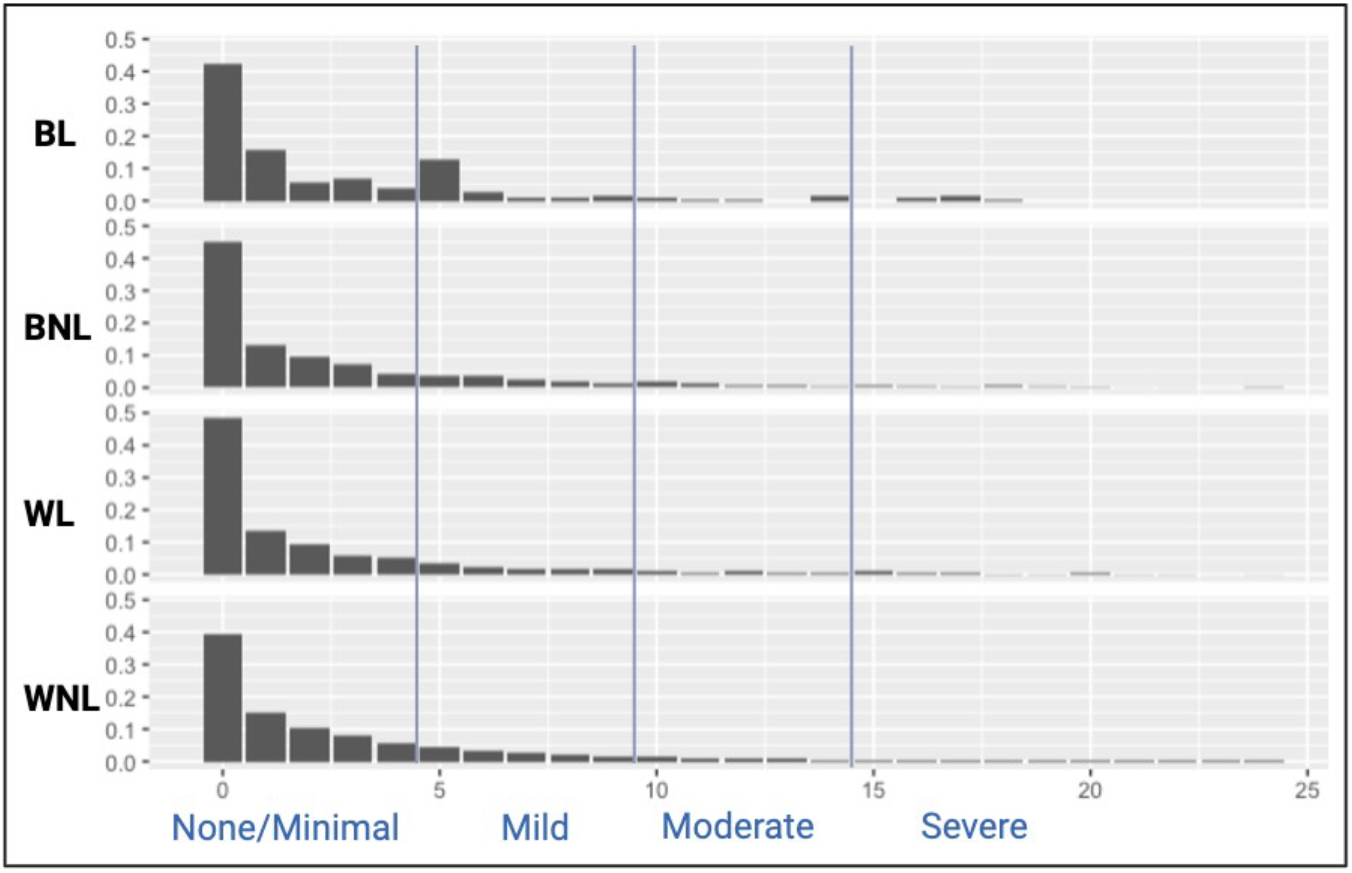
Survey-weighted density distributions of PHQ-8 scores and scale categorization by race/ethnicity. BL, Black-Latiné; BNL, Black Non-Latiné; WL, White-Latiné; WNL, White Non-Latiné. Vertical lines mark scale category cutoffs.

## Discussion

The purpose of this study is to identify sociodemographic differences between Black-Latinés compared to White-Latinés, Non-Latiné Blacks, and Non-Latiné Whites in the U.S. and to assess the prevalence of depression and anxiety in Black-Latinés compared to White-Latinés, Non-Latiné Blacks, and Non-Latiné Whites counterparts. Overall, Black-Latinés were more likely than both White-Latinés and Non-Latiné Blacks to self-report more severe symptoms of anxiety and depression and to have a clinical history of depression and anxiety. Currently, there is a dearth of literature exploring the mental health needs of Black-Latiné. Studies argue that Non-Latiné Blacks may have lower rates of depression compared to Non-Latiné Whites due to misdiagnoses, the resilient nature of Non-Latiné Blacks, and because of strong community ties [28,29]. However, very little is known about how Black-Latinés fair compared to not only their Non-Latiné White counterparts but to White-Latinés and Non-Latiné Blacks. Our findings reveal there may be mental health disparities not only between racial groups as evidenced by Black-Latinés having a higher prevalence of poor mental health symptoms than White-Latinés, but also within racial groups as Black-Latinés have a higher prevalence of poor mental health symptoms than Non-Latiné Blacks.

Although Black-Latinés are grouped under the “Black” racial category, Black-Latinés within our sample are different with regard to sociodemographic factors and mental health symptoms.

Previous literature corroborates the need to study subgroups within the broader Black racial category [30,31]. Williams et al. previously found that Black Caribbean men had a higher risk for psychiatric disorders compared to African American men [31]. Furthermore, immigration history and acculturation within the U.S. influenced the incidence of psychiatric disorders with third generation Black Caribbean men having higher rates of psychiatric disorders compared to first-generation immigrants [30]. In our study, even though Black-Latinés were more likely to report being newer immigrants they still report more severe anxiety and depression symptoms than both White-Latinés and Non-Latiné Blacks. This leaves unclear how race, ethnicity, and acculturation intersect to influence mental health within this population.

Racial/ethnic minorities often face significant challenges in accessing mental health treatment and despite insurance status and insurance type, Non-Latiné Whites are more likely to receive higher-quality care [32]. Insurance status alone cannot explain disparities in mental health. In our study, even though Black-Latinés were more likely to report having health insurance than both White-Latinés and Non-Latiné Blacks and thus were more likely to report a clinical diagnosis of anxiety and depression, it is unclear if this led to treatment. A study by Nelson et al. found that racial/ethnic minorities are more likely to refuse medical treatment compared to their White counterparts due to a variety of reasons including discrimination, language barriers, and health information miscommunication [33,35].

### Strengths and Limitations

The study has several limitations. First, this is a cross-sectional analysis where we cannot make any assumptions about temporality and causality. Second, depression and anxiety symptoms were self-reported. Although respondents were asked if they had a history of being diagnosed with depression and anxiety, there is still potential for misclassification. Individuals influenced by cultural and social desirability are more likely to underreport symptoms due to the stigma associated with mental health [7,8]. Third, the NHIS excluded individuals without a permanent address, unhoused, wards of the state, resides in US territories, active-duty military personnel, U.S nationals living abroad or bases, and long-term care facilities [12,15]. This may result in undersampling of groups that may be potentially at higher risk for poor mental health thus leading to an undercount of the prevalence of depression and anxiety in our sample. For instance, Black individuals are more likely to be institutionalized than their White counterparts [34]. Finally, the GAD-7 and PHQ-8 has not been culturally validated in our populations of interest [35]. A culturally biased screening tool may result in scores that do not truly reflect the mental health status of racial/ethnic individuals.

This study has several strengths. First, the NHIS dataset is a large nationally representative sample of non-institutionalized individuals. Second, historically, the NHIS has oversampled for Black and Latiné populations and made sampling redesigns to increase the precisions of estimates for both groups [2,5]. Oversampling of subgroups enabled us to compare the mental health outcomes of disaggregated racial/ethnic groups.

## Conclusion

The findings of this descriptive paper indicate that Black-Latinés are potentially experiencing mental health that is worse than other racial/ethnic groups, particularly White-Latinés and Non-Latiné Blacks. Previous studies have aggregated Black-Latiné data with Non-Latiné Black data potentially masking disparities within Black populations. The U.S. is projected to become a majority-minority country by 2044 [16]. Studies often collapse Black-Latinés into the Black racial category without taking into consideration ethnic identity, different immigration histories, language barriers, and cultural factors that distinguish Black subgroups. To ameliorate any potential poor mental health in Black-Latinés, intersectional policies that consider both race and ethnicity and address the impacts of immigration and acculturation should be generated and enacted. Future studies should explore the mechanism between multiple intersectional identities and poor mental health.

## Data Availability

All data produced are available online at https://www.cdc.gov/nchs/nhis/documentation/index.html

## Declarations

### Data Availability

The data that support the findings of this study are available from the National Health Interview Survey at https://www.cdc.gov/nchs/nhis/documentation/2022-nhis.html.

### Ethics Approval

This research was reviewed by the UCSF IRB and was granted exemption under Category 4: secondary research for which consent is not required. **IRB Number:** 25-44668

